# Case Fatality Risk of Norovirus in England During a Period of Strain Replacement, 2022/23 - 2024/25 Seasons

**DOI:** 10.64898/2025.12.19.25342373

**Authors:** Maria L. Tang, Amy Douglas, Cristina Celma, Roberto Vivancos, Gauri Godbole, Thomas Ward, Jonathon Mellor

## Abstract

**Background:** Norovirus causes substantial burden to healthcare systems. England experienced high activity in recent seasons alongside a shift in the dominant genotype from GII.4 to GII.17. It remains unclear whether this increased burden reflects changes in severity or other transmission mechanisms associated with the strain replacement.

**Methods:** Individual-level testing and mortality data in England from 2022/23-2024/25 seasons were linked from national surveillance systems. Piece-wise exponential additive mixed models estimated all-cause case-fatality risk (CFR) for death within 28 days of a positive test, adjusting for age, location, primary or secondary care case identification and temporal effects. Additional analysis tested the effect of the death linkage threshold.

**Results:** CFR did not differ significantly between GII.4 and GII.17, but increased markedly with age and was substantially higher for cases detected in secondary versus primary care. At the 2024/25 season peak, the CFR for the average cohort case (age 75) was 8.01% (95% CI: 6.88–9.33%) for cases identified in secondary care compared with 1.18% (95% CI: 0.61–2.25%) for primary care, likely driven by different case mixes and unmeasured confounding between patients managed in these settings.

**Interpretation:** We found no evidence that the newly dominant GII.17 genotype had higher CFR than GII.4 among test-positive cases. The increased burden is therefore more consistent with changes in transmissibility, population immunity, or testing dynamics than increased genotype-specific severity. Norovirus remains a major public health threat, and understanding genotype switching supports enhanced surveillance, expanded genotyping, routine clinical data linkage and advanced modelling to inform control strategies.

## Introduction

Norovirus is a highly contagious virus causing diarrhoea and vomiting symptoms, with vulnerable groups at increased risk of severe or prolonged illness requiring hospital care. While not inherently severe, its high transmissibility in closed settings adversely impacts at-risk populations, particularly the elderly and hospital patients [1]. Community infections are substantially under-ascertained, as national surveillance mostly detects outbreaks in care homes or hospital wards, biasing data toward more severe cases [2]. For each reported case in the UK, an estimated 288 community infections occur [3], possibly exacerbated by guidance to manage symptoms at home. Norovirus incidence peaks in winter, increasing pressure on healthcare providers through outbreaks in closed settings and its impact on vulnerable populations. This is particularly due to bed-blocking closures for infection control and prevention, in addition to staff sickness and wider system disruption [4].

Noroviruses are classified into 10 genogroups (GI-GX) and further divided into 49 genotypes [5], with limited cross-immunity between genotypes [6]. Multiple norovirus genotypes co-circulate globally, however GII.4 is the most commonly detected, with the GII.4/Sydney/2012 strain dominating most of the last decade [7] [8]. Historically, dominant GII.4 strains are periodically replaced over semi-regular cycles [9]. During the 2023/24 season, surveillance in multiple countries including England detected the re-emergence of GII.17, which unusually replaced GII.4 as the dominant genotype [10]. High norovirus activity continued into 2024/25, with England seeing its highest ever levels of laboratory reports, driven by co-circulation following the re-emergence of GII.4/Sydney/2012 [11] [12].

With the re-emergence of genotype GII.17, there was a substantial increase in norovirus activity in England over winter 2024/25 [13]. Drivers for this could include increased severity, higher transmissibility, or lower population immunity to GII.17 compared to GII.4, or a combination of these factors. Untangling these effects is challenging [14], particularly when analyses rely on aggregate data [15].

Previous studies have compared severity and transmissibility between norovirus genotypes, but none yet for the recent GII.17 strain. Seasons dominated by GII.4 have shown higher hospitalisation and mortality rates than years with dominant non-GII.4 genotypes [16] [17]. However, during the emergence of a new GII.4 variant in England in 2002/03, the relative mortality risk among elderly individuals was lower than in other seasons [18]. Overall, the study estimated 80 norovirus-attributable deaths per year in individuals aged 65 years or older in England. Other studies have estimated norovirus severity but not by genotypes. Among US veterans, norovirus was associated with a slight increase in hospitalisation risk and a larger increase in mortality risk that decreased over time [19].

Case-fatality risk (CFR), the probability that a reported case results in death, is a key measure of disease severity. CFR in studies can vary widely due to differences in age, comorbidities and setting (e.g. hospital, residential care) [1] [20], and whether all-cause or norovirus-attributable deaths are used. A review of 52 international studies found high heterogeneity in CFR, with an overall model-based CFR of 0.025% (95% CI: 0.003%, 0.195%) [21]. CFR from seven studies of residents in long-term care facilities ranged from 0.3% to 1.6% [22]. Most previous work relies on aggregate data or data from specific subpopulations, which makes national-level inferences more difficult.

In this study, we use linked individual-level data encompassing positive laboratory test results, genotyping data, hospital admissions and deaths to explore the changing norovirus genotype dominance across three seasons between 2022 and 2025 in England. By linking genotyping data with mortality records, we estimate genotype-specific case-fatality risk, giving a measure of relative severity across strains.

## Methods

### Data sources

This retrospective cohort study included individuals with a positive norovirus test in England. We combined routinely collected individual-level data to assess severity, linking laboratory test results to mortality and hospitalisation records.

### Genotyping data

Genotyping data in England came from the UK Health Security Agency’s (UKHSA) Modular Open Laboratory Information System (MOLIS) [23]. The Enteric Virus Reference Unit genotype a subset of norovirus-positive specimens for surveillance. Most specimens were referred by the seven regional laboratories, covering the nine English regions (with single laboratories serving both the West and East Midlands, and London and the East of England). Guidance at the time requested laboratories sent up to three norovirus-positive specimens associated with any one hospital or community outbreak, and up to five further randomly selected non-outbreak specimens each week, prioritising specimens with the highest viral load based on PCR cycle threshold values. Fewer specimens came from local National Health Service (NHS) laboratories which were encouraged to refer specimens for genotyping during winter 2024/25 in response to the rise in GII.17, selecting up to two specimens per week with the highest viral load. We de-duplicated (see Supplementary Section 1) and analysed genotyping data with specimen date from 1 July 2022 to 31 March 2025, before statutory notifiable reporting of norovirus test results began on 6 April 2025 [24].

### SGSS data

Diagnostic laboratories in England have a statutory duty to report positive detection of notifiable organisms and do this electronically, through UKHSA’s Second Generation Surveillance Service (SGSS) [23]. We extracted positive norovirus test results from England from SGSS, which captures a larger set of tests more representative of overall burden but lacks genotyping. SGSS is one of the three systems used by UKHSA to monitor epidemiological trends for norovirus [25]. SGSS also contains more demographic and contextual information, including patient residential geography and the healthcare level that requested the test, such as general practice (GP), hospital inpatient, hospital outpatient and accident & emergency (A&E). We de-duplicated (see Supplementary Section 1) and analysed records with specimen date from 1 July 2022 to 31 March 2025.

### Severe outcomes

All-cause death registrations were obtained from the Office of National Statistics (ONS) for dates of death up to 23 July 2025 [26]. Hospitalisation data is detailed in Supplementary Section 2.

### Data linkage

Records were linked using NHS number (unique patient identifier) and relevant dates:

- Deaths were separately attributed to infection episodes from the MOLIS and SGSS data by linking each death to any positive result occurring within 28 days prior to the date of death. When multiple test results met this criterion, the most recent test result was selected, assuming more recent infection episodes are more attributable to deaths. Sensitivity of results on the 28-day death threshold is detailed in the supplementary material.
- To link MOLIS and SGSS test results, each MOLIS test result was linked to the closest SGSS test result within a 21-day window either side, according to specimen dates. Some MOLIS test results from regional laboratories did not have corresponding SGSS entries. Most linked MOLIS-SGSS pairs shared identical specimen dates; for those that did not, the earliest specimen date of the two was used in the following analysis.
- We refer to the datasets of de-duplicated MOLIS, SGSS and MOLIS, and SGSS test results and linked deaths as **MOLIS-deaths**, **MOLIS-SGSS-deaths** and **SGSS-deaths** data respectively.
- Hospitalisations were analogously linked to positive tests (detailed in Supplementary Section 2).

### Causal structure

Causal relationships contributing to death of norovirus cases, whether attributable to norovirus or not, are shown in Figure 2. Observed data include age and some healthcare characteristics (level of healthcare, region), but other patient demographics, healthcare characteristics (e.g. specialism and size) and comorbidities that affect risk of death are unobserved. Healthcare system pressure can also increase mortality risk due to resource constraints – although unobserved, temporal variation can be partially adjusted for by including calendar time in our model.

**Figure 1:**
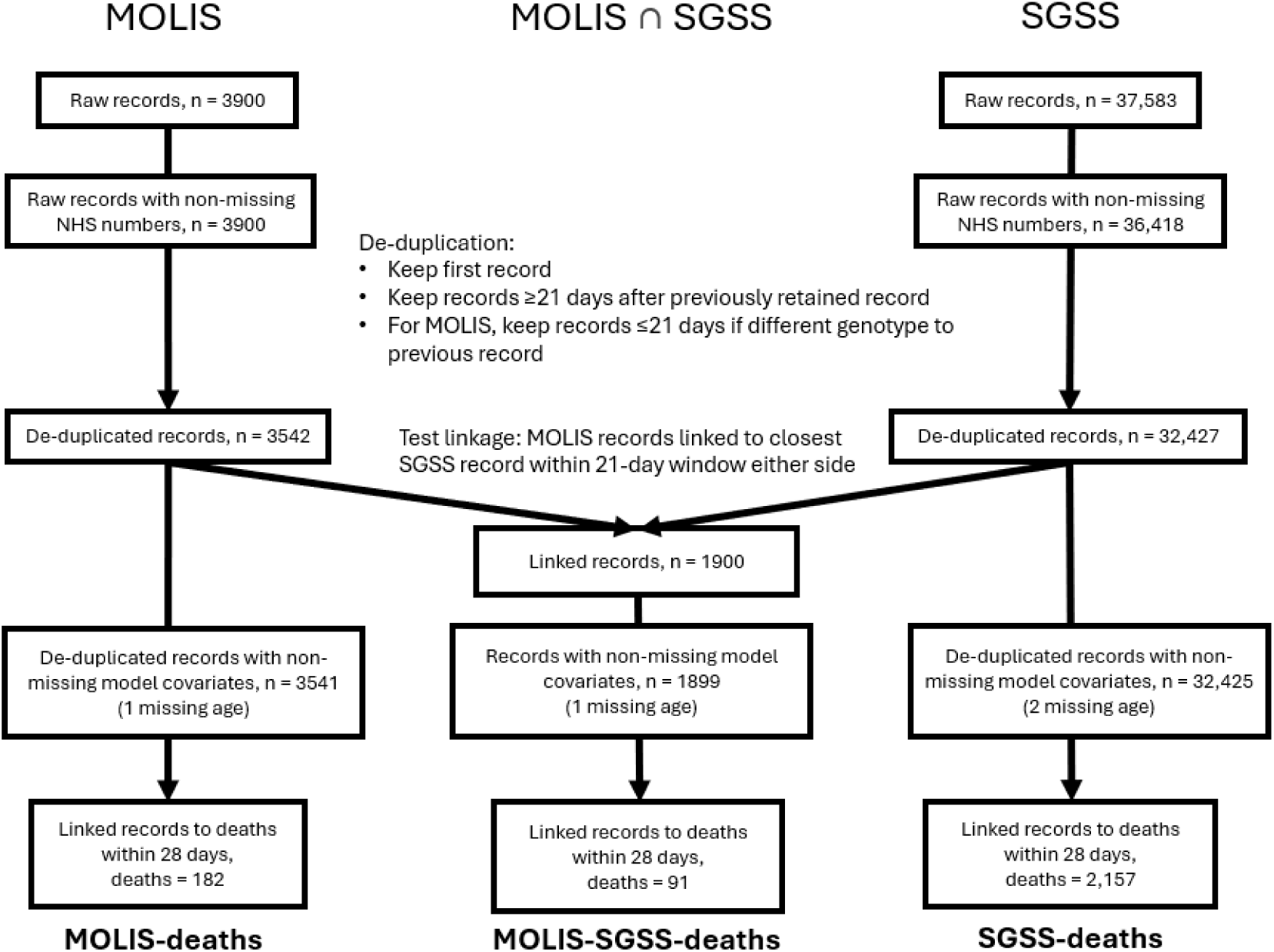
Data flow from raw data sources to de-duplicated linked test result records and deaths that are used in the case-fatality models.

**Figure 2.**
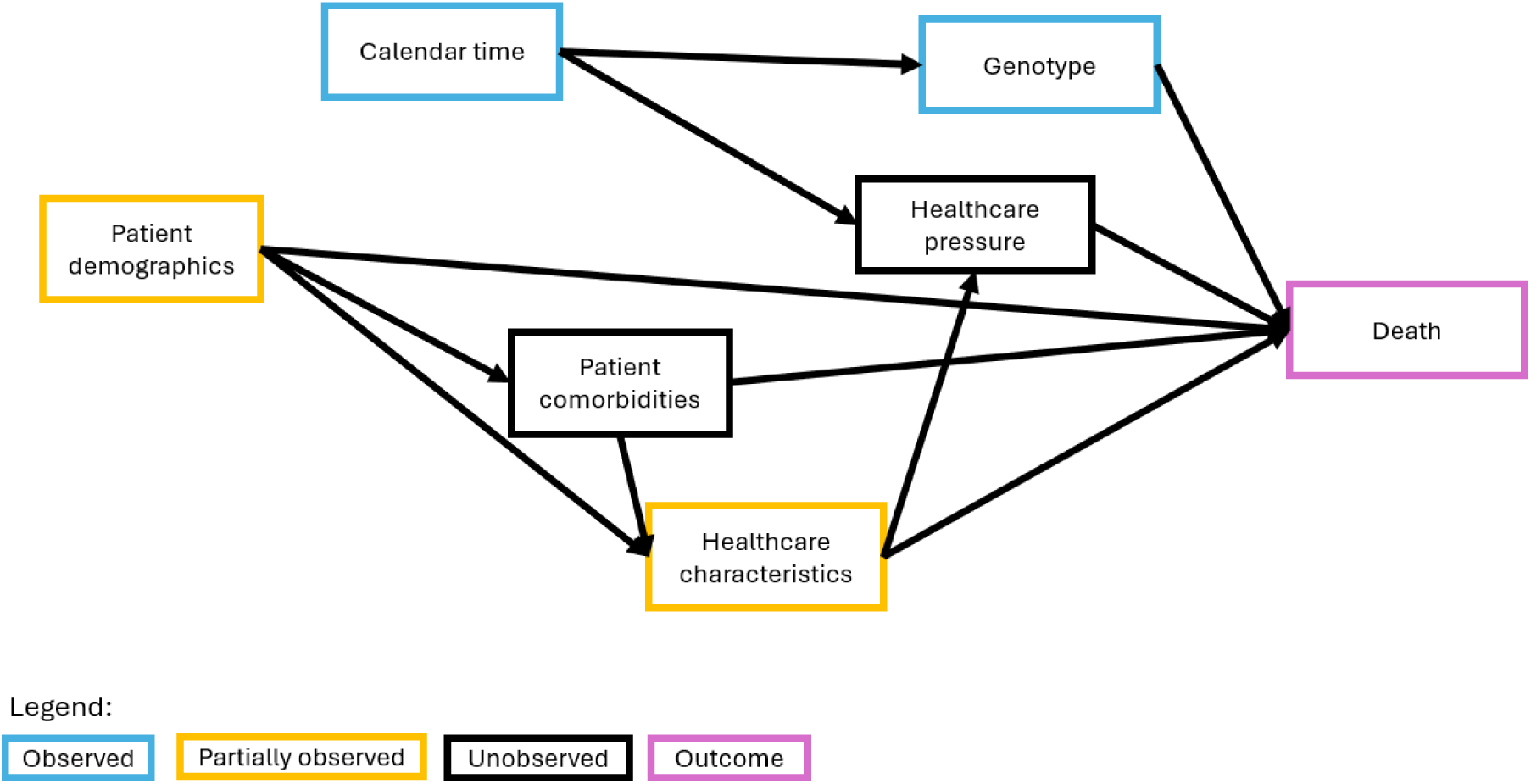
Directed acyclic graph (DAG) of causal effects contributing to the death of a detected norovirus case, whether attributable to norovirus or not. Observed effects are in blue, partially observed effects in orange, and the outcome (death) in purple. Arrows show the direction of causal relationships.

Not all individuals with norovirus have the same probability of being tested, introducing selection bias (detailed in Supplementary Section 3). Observed severity among the cohort of ascertained cases is therefore not representative of severity across all infections.

### Case-fatality model

Piece-wise exponential additive mixed models (PAMMs) were used to estimate hazard ratios (HRs) and risk of death following a positive norovirus test. PAMMs give a flexible framework to model time-to-event data with non-linear and time-varying effects using techniques from generalised additive mixed models (GAMMs) [27]. These were fit using the R packages “mgcv” [28] and “pammtools” [29].

Models were fit separately to MOLIS-deaths (genotyped), SGSS-deaths (larger sample size including additional covariates, but no genotype data) and their intersection, MOLIS-SGSS-deaths data. Some individuals had multiple test results. Test results were followed up until the earliest of death, 28 days after specimen date, the specimen date of the next test result of that individual, or the data cutoff date. When follow-up ended on or before the specimen date, follow-up time was set to 1 day. Follow-up was split into daily intervals from specimen date for MOLIS-deaths and MOLIS-SGSS-deaths, reflecting the daily resolution of specimen and death dates. For SGSS-deaths models, two-day intervals were used to reduce computational load. Covariates were included for genotype, healthcare level and lab region. Groupings for these are given in Supplementary Section 4.

Based on the DAG (Figure 2) and subject matter expert opinion elicitation, we specified the following hazard functions *h*_*i*_(*t*) for person-record-episode *i* at time *t* (time since specimen date):

MOLIS-deaths:

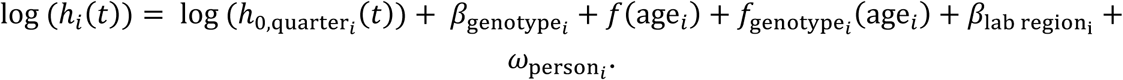

MOLIS-SGSS-deaths:

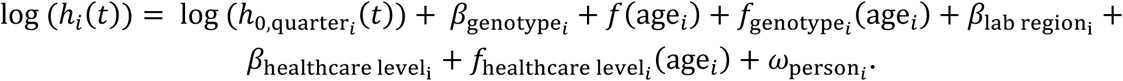

SGSS-deaths:

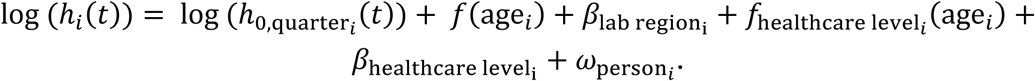

Here, log (*h*_0,quarter*i*_(*t*)) are stratum-specific log baseline hazards for specimen date quarter. This captures potential temporal changes in strain virulence, healthcare-seeking behaviour, healthcare access, or hospital pressures that could influence severity. Specimen date quarter is used due to the sparse data in some months in the MOLIS-deaths and MOLIS-SGSS data, and also used for SGSS-deaths models for consistency. *β* are fixed effects, *f*(age_*i*_) are global splines fit to age, *f*_genotype*i*_(age_*i*_) and *f*_healthcare level*i*_(age_*i*_) are age splines specific to genotypes and healthcare levels, and *ω*_person*i*_ are person-level random effects to account for multiple record-episodes per individual. We include genotype and healthcare-level-specific age effects to allow age-related severity patterns to differ across groups.

## Results

Between 1 July 2022 and 31 March 2025, England experienced seasonal waves of norovirus across cases and hospitalisations, beginning around October and peaking in spring (Figure 3). The 2022/23 season was dominated by GII.4, but midway through the 2023/24 season, GII.17 became the dominant genotype. The 2023/24 season was characterised by a flat peak over many months. The 2024/25 season experienced a higher peak in positive test results, with GII.17 continuing to dominate. GII.4 re-emerged in early 2025 and the co-circulation of these two genotypes resulted in a higher peak. These seasonal peaks are also seen in the severe outcomes, but less apparent in the smaller sample size of the MOLIS genotyping data.

**Figure 3:**
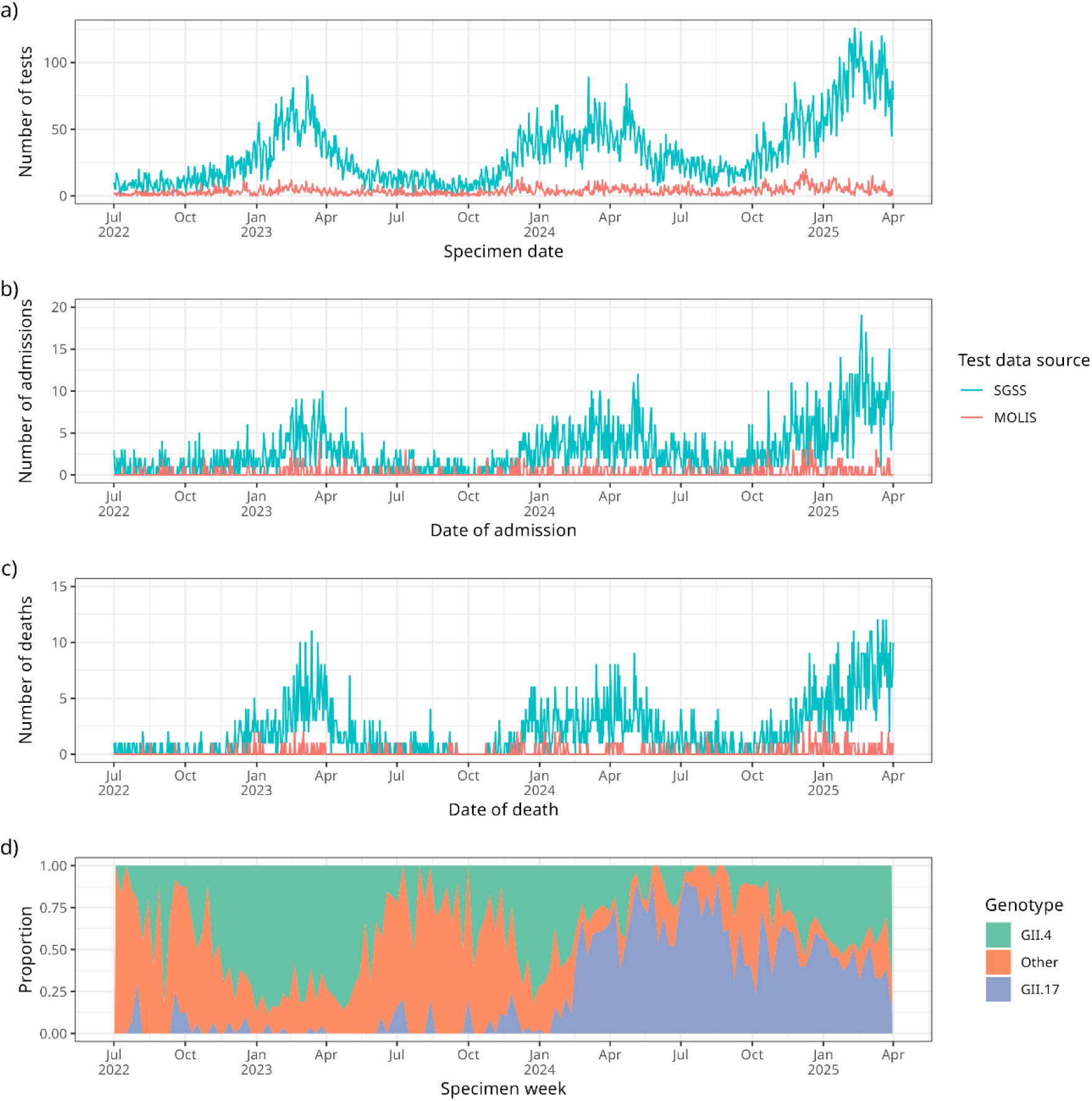
For **SGSS-deaths** and **MOLIS-deaths** linked datasets, a) counts of de-duplicated positive norovirus test results over time, b) counts of hospital admissions within 14 days after a linked positive norovirus test result, c) counts of deaths within 28 days after a linked positive norovirus test result. For MOLIS tests, d) genotype proportion of positive norovirus test results.

Positive test results are concentrated in the elderly and infants, with MOLIS having more of a bias towards infants (Supplementary Figure 2). Deaths are concentrated in the elderly, with some deaths also observed in infants. In MOLIS cases with a linked death, most deaths are seen within 60 days (Supplementary Figure 3, Supplementary Table 1). Based on raw counts, the unadjusted case-fatality risk is highest for GII.4, lower for GII.17, and much lower for other genotypes (Table 1). The unadjusted case-fatality risk by age is highest for the elderly and smallest for infants. For MOLIS cases with a linked hospitalisation, tests are mostly taken after admission (Supplementary Figure 3).

**Table 1:** Unadjusted raw counts of de-duplicated positive test results from **MOLIS-deaths** data within 28 days.

| Covariate | Positive tests | Deaths | Deaths / positive tests |
| --- | --- | --- | --- |
| **Overall** |  |  |  |
|  | 3542 | 182 | 0.051 |
| **Genotype** |  |  |  |
| GII.4 | 1297 | 92 | 0.071 |
| GII.17 | 1019 | 53 | 0.052 |
| Other | 858 | 13 | 0.015 |
| Unknown | 368 | 24 | 0.065 |
| **Age group** |  |  |  |
| 0-4 | 864 | 3 | 0.003 |
| 5-19 | 183 | 2 | 0.011 |
| 20-49 | 355 | 6 | 0.017 |
| 50-74 | 708 | 38 | 0.054 |
| 75+ | 1431 | 132 | 0.092 |
| **Region** |  |  |  |
| East of England | 332 | 18 | 0.054 |
| London | 48 | 1 | 0.021 |
| Midlands | 301 | 21 | 0.070 |
| North East and Yorkshire | 476 | 19 | 0.040 |
| North West | 1609 | 83 | 0.052 |
| South East | 82 | 4 | 0.049 |
| South West | 694 | 36 | 0.052 |

During the peak of the 2024/25 season, the average case in the MOLIS-deaths cohort (median age = 64, modal region = North West) had a case-fatality risk (CFR) of 6.99% (95% CI: 3.88%, 12.53%) for genotype GII.4, and 5.04% (95% CI: 2.70%, 9.38%) for genotype GII.17, for death within 28 days of a positive norovirus test result (Table 2). CFR in the 2022/23 and 2023/24 seasons were similar. CFR increased monotonically by age, and is similar between the two genotypes with largely overlapping confidence intervals (Figure 4). No significant difference was observed in the estimated risk of death between individuals infected with GII.4 and GII.17 (Supplementary Figure 4), but there were some differences in risk between individuals with specimens sequenced in different regions.

**Figure 4:**
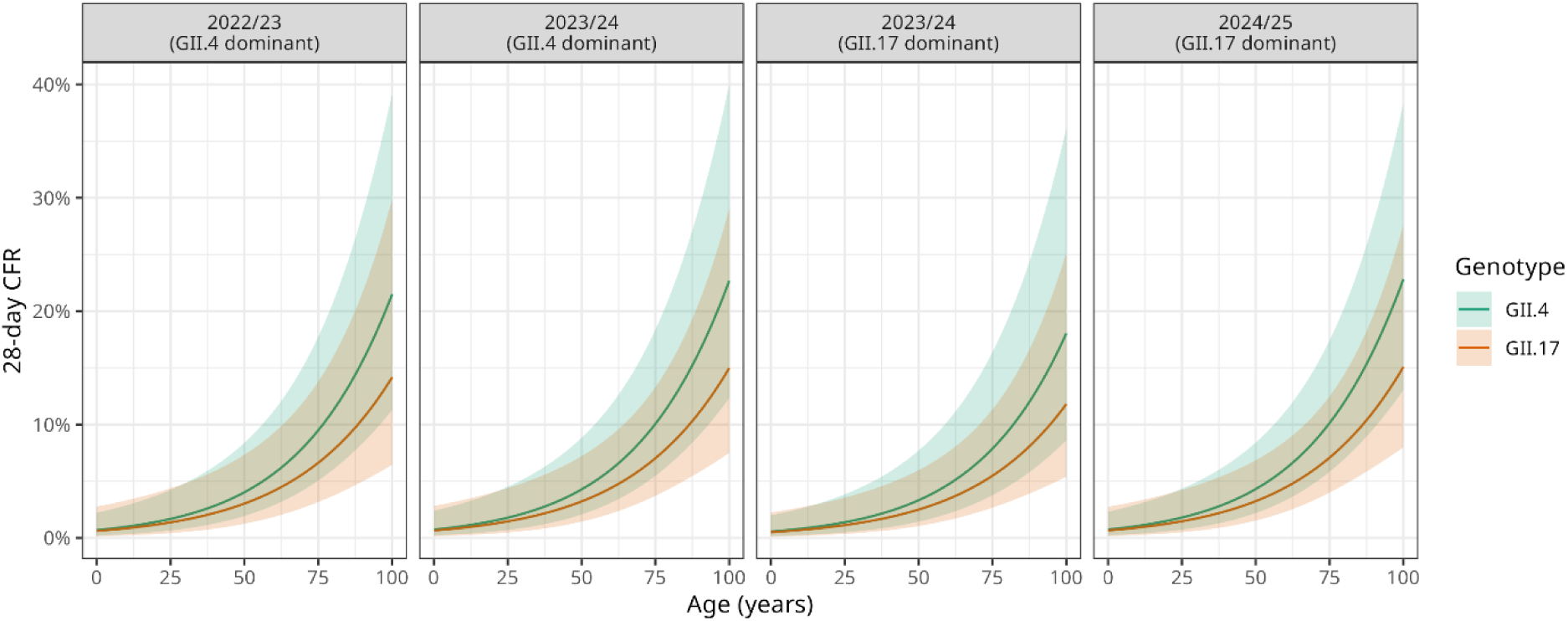
Predicted age-varying case fatality risk (CFR) within 28 days of a positive test with 95% confidence intervals by genotype and time period using **MOLIS-deaths** data, for tests taken in the modal region (North West). Time period facets were chosen to reflect periods of peak activity in each season. Two time periods are shown for the 2023/24 season to reflect periods of GII.4 or GII.17 dominance. In order, the time periods refer to test results with specimen date in the calendar quarter beginning January 2023, January 2024, April 2024 and January 2025.

**Table 2:** Case-fatality risk by genotype and time period of death within 28 days of a positive norovirus test for an average person in the MOLIS-deaths cohort (median age = 64, modal region = North West). Time period facets were chosen to reflect periods of peak activity in each season. Two time periods are shown for the 2023/24 season to reflect periods of GII.4 or GII.17 dominance. In order, the time periods refer to test results with specimen date in the calendar quarter beginning January 2023, January 2024, April 2024 and January 2025.

| Genotype | CFR % (95% CI) |
| --- | --- |
| **2022/23 (GII.4 dominant)** |  |
| GII.4 | 6.54 (3.39, 12.66) |
| GII.17 | 4.71 (2.16, 10.27) |
| **2023/24 (GII.4 dominant)** |  |
| GII.4 | 6.94 (3.64, 13.20) |
| GII.17 | 5.00 (2.52, 9.95) |
| **2023/24 (GII.17 dominant)** |  |
| GII.4 | 5.41 (2.51, 11.68) |
| GII.17 | 3.89 (1.82, 8.39) |
| **2024/25 (GII.17 dominant)** |  |
| GII.4 | 6.99 (3.88, 12.53) |
| GII.17 | 5.04 (2.70, 9.38) |

Table 3 shows the CFR for the average case in the SGSS-deaths cohort (median age = 75, modal region = South West) by healthcare level and time period. During the peak of the 2024/25 season, the CFR was 8.01% (95% CI: 6.88%, 9.33%) for cases identified in secondary care, and 1.18% (95% CI: 0.61%, 2.25%) for those identified in primary care. CFR in the 2022/23 and 2023/24 seasons were similar. CFR mostly increased monotonically by age for all healthcare levels (Figure 5). The risk of death was significantly higher for cases identified in secondary care compared to those in primary care for all ages up to 97 (hazard ratio at median age 0.14 (95% CI: 0.07, 0.27)) and for those identified in other types of healthcare for ages 18 and above (hazard ratio at median age 0.62 (95% CI: 0.44, 0.88)) (Supplementary Figure 5). There were some significant differences in risk between individuals tested in different regions.

**Figure 5:**
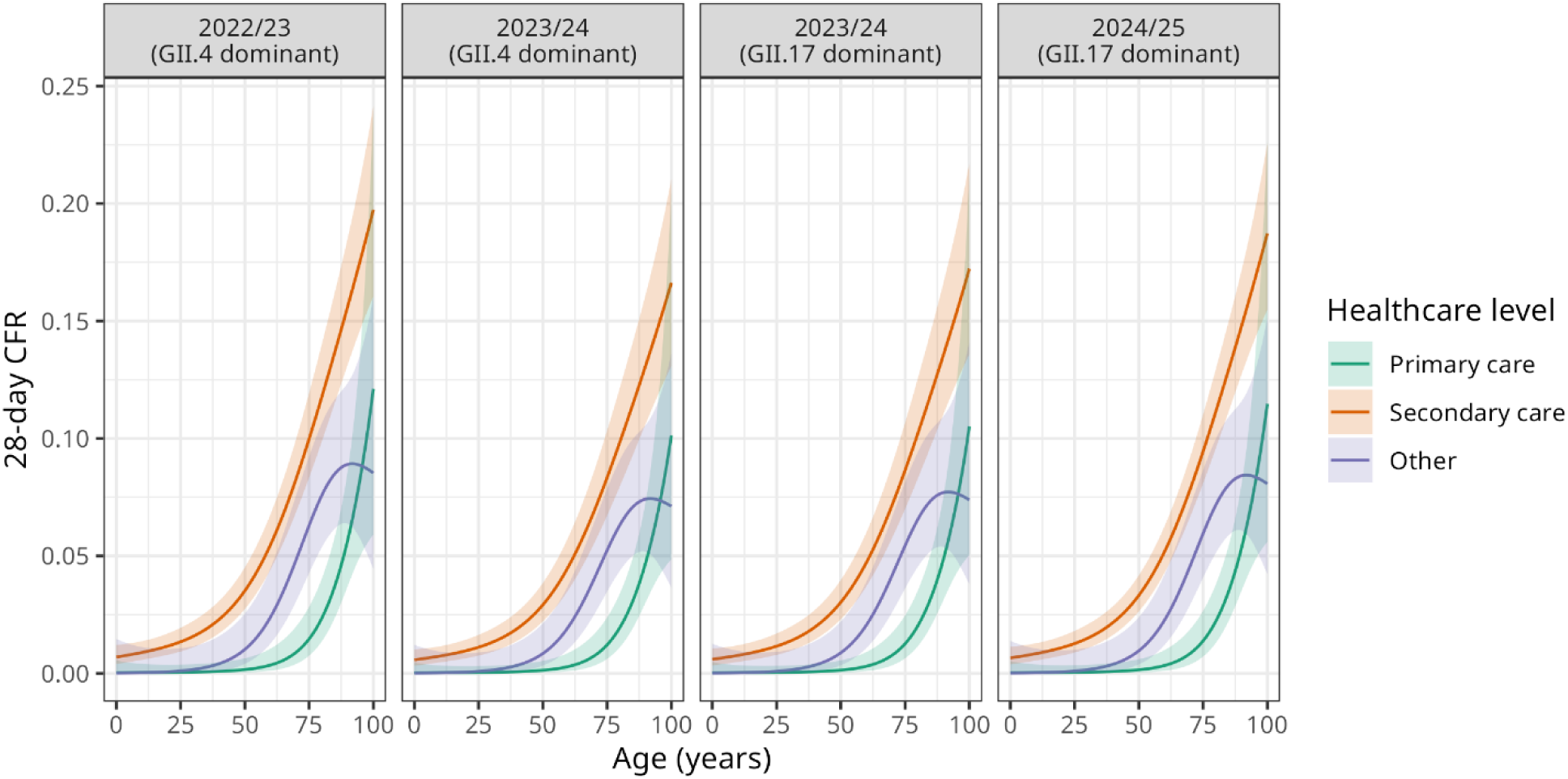
Predicted age-varying case fatality risk (CFR) within 28 days of a positive test with 95% confidence intervals by healthcare level and time period using **SGSS-deaths** data. Time period facets were chosen to reflect periods of peak activity in each season. Two time periods are shown for the 2023/24 season to reflect periods of GII.4 or GII.17 dominance. In order, the time periods refer to test results with specimen date in the calendar quarter beginning January 2023, January 2024, April 2024 and January 2025.

**Table 3:** Case-fatality risk of death within 28 days of a positive norovirus test for an average case in the SGSS-deaths cohort (median age = 75, modal region = South West) identified in primary or secondary care. Time period facets were chosen to reflect periods of peak activity in each season. Two time periods are shown for the 2023/24 season to reflect periods of GII.4 or GII.17 dominance. In order, the time periods refer to test results with specimen date in the calendar quarter beginning January 2023, January 2024, April 2024 and January 2025.

| **Healthcare level** | **CFR % (95% CI)** |
| --- | --- |
| **2022/23 (GII.4 dominant)** |  |
| Primary care | 1.25 (0.65, 2.40) |
| Secondary care | 8.47 (7.13, 10.06) |
| **2023/24 (GII.4 dominant)** |  |
| Primary care | 1.03 (0.53, 2.01) |
| Secondary care | 7.06 (5.70, 8.75) |
| **2023/24 (GII.17 dominant)** |  |
| Primary care | 1.07 (0.55, 2.09) |
| Secondary care | 7.33 (5.93, 9.07) |
| **2024/25 (GII.17 dominant)** |  |
| Primary care | 1.18 (0.61, 2.25) |
| Secondary care | 8.01 (6.88, 9.33) |

Genotype and healthcare level (primary vs secondary) effects modelled from the smaller MOLIS-SGSS-deaths data were statistically insignificant (Supplementary Section 5). For all datasets, results were similar when instead using a 14-day or 60-day threshold for associating deaths with a positive norovirus test (Supplementary Section 6).

## Discussion

Norovirus imposes a substantial burden on the population and healthcare system in England, costing the NHS over £100 million annually [30]. This was particularly acute during the 2023/24 and 2024/25 seasons, coinciding with a change in the dominant genotype from GII.4 to GII.17. Differences in infection dynamics relating to the genotype shift may have contributed to the observed increase in norovirus activity. By estimating genotype-specific case-fatality risk (CFR), this study found no evidence of increased mortality of GII.17 compared to GII.4. The increased activity is therefore more likely driven by changes in transmissibility, population immunity, or testing dynamics than an increase in genotype-specific severity.

There was no evidence of a difference in CFR between genotypes GII.4 and GII.17. However, CFR was substantially higher for cases identified in secondary care compared with primary care. During the 2024/25 peak, the CFR for the average SGSS patient (age 75) was estimated as 8.01% (95% CI: 6.88%, 9.33%) for cases identified in secondary care, and 1.18% (95% CI: 0.61%, 2.25%) in primary care, for deaths within 28 days of a positive norovirus test result. This is the 28-day all-cause mortality risk among test-positive norovirus cases, and so likely overestimates norovirus-attributable death. Attributing deaths to norovirus is difficult, as severe illness occurs primarily in older adults with comorbidities [19] [22]. These patients are also more often in hospitals or care homes where testing is likely and have higher viral loads which are more likely to be referred for genotyping, introducing selection bias.

Direct comparison with CFRs from other studies is challenging due to different cohorts, surveillance capability and death linkage methods. Our estimated cohort CFR is much larger than a model-based CFR of 0.025% (95% CI: 0.003%, 0.195%) from 52 international studies, many of which had small sample sizes and no deaths [21]. Some studies used norovirus diagnostic codes to attribute deaths, giving a smaller CFR than all-cause mortality risk. As our results suggest mortality risk greatly increases with age, reported CFRs depend substantially on the cohort’s age structure, which may explain regional differences seen in our CFR.

The absence of a detectable difference in CFR between GII.4 and GII.17 suggests that the 2024/25 wave of severe outcomes was unlikely driven by increased severity of GII.17. Instead, increased transmissibility or reduced population immunity for the new dominant GII.17 genotype likely contributed, consistent with limited population exposure to GII.17 after almost a decade of GII.4 dominance until 2023 [6] [10]. This research is timely given current ongoing development of norovirus mRNA vaccines [31]as the choice of vaccine strain composition and evaluation strategy should consider genotype-specific severity. Meanwhile, if GII.17 is not inherently more severe, public health actions should continue to prioritise hygiene and cleaning in community and healthcare settings.

Use of individual-level mortality records and test results enabled estimation of genotype-specific risk while adjusting for patient and healthcare provider characteristics such as age and location, offering advantages over aggregate CFR approaches. Linkage of test and death data allowed assessment of linkage definitions, showing robustness of hazard ratio results (Supplementary Section 6). PAMMs provided flexibility in model structure while accounting for right censoring. However, we were unable to adjust for all confounding factors that can bias our results [32], such as patient comorbidities. Sample size constraints also limited power in analyses when adjusting for both genotype and healthcare level.

After this study’s period, norovirus became notifiable in England (6 April 2025), requiring laboratories to report positive results to UKHSA. This will improve completeness of test data and may partially enhance case ascertainment. However, testing will likely remain biased towards more severe cases, inflating CFR estimates of ascertained cases compared to all cases [32]. Further, the change in reporting could shift testing practice unpredictably. Expanded routine genotyping, including outside health and social care settings, would improve estimates of genotype-specific severity and allow for adjustment for confounding. Current prioritisation of high-viral-load specimens for genotyping is another bias towards severe cases but is operationally necessary to ensure genotyping is possible.

This study focused on case-fatality risk, but hospitalisation is another important outcome. However, from linked case-hospitalisation data, most tests were taken after hospital admission. While tests taken shortly after admission may indicate community-acquired infections which are then hospitalised, the longer delays observed for most cases are more consistent with nosocomial transmission. Hence, this data would not be so appropriate for estimating case-hospitalisation risk as most cases were already in hospital prior to contracting norovirus. Nosocomial transmission also disproportionately affects higher risk patients (elderly or comorbid) and hence raises observed CFR values [33]. Community studies could improve understanding of severity by estimating infection-hospitalisation and infection-fatality risks [34].

Future research can investigate how changes in transmissibility, immunity and susceptibility jointly drive genotype replacement events. Further mechanistic transmission modelling of norovirus would help clarify how changes in genotype dominance translate into observed epidemiological outcomes [35]. Strengthening routine linkage and surveillance of genomic, clinical and demographic data will be crucial for assessing the impact of genotype shifts and informing vaccine development.

## Acknowledgements

We thank UKHSA data operations colleagues for their work in the access, processing and maintenance of the data used in this paper.

## Author contributions

Maria L. Tang: Conceptualisation; Methodology; Software; Formal analysis; Writing – Original Draft; Writing - Review & Editing; Visualization.

Amy Douglas: Conceptualisation; Methodology; Writing - Review & Editing; Supervision. Cristina Celma: Resources, Data Curation, Writing - Review & Editing.

Roberto Vivancos: Methodology; Writing - Review & Editing. Gauri Godbole: Writing - Review & Editing.

Thomas Ward: Methodology; Validation; Writing - Review & Editing.

Jonathon Mellor: Conceptualisation; Methodology; Software; Validation; Writing - Original Draft; Writing - Review & Editing; Supervision.

## Data availability

Synthetic data and the code for the paper can be found on GitHub: https://github.com/maria-tang/norovirus-case-fatality

An application for data access can be made to the UK Health Security Agency. UKHSA operates a robust governance process for applying to access protected data that considers:

- the benefits and risks of how the data will be used
- compliance with policy, regulatory and ethical obligations
- data minimisation
- how the confidentiality, integrity, and availability will be maintained
- retention, archival, and disposal requirements
- best practice for protecting data, including the application of ‘privacy by design and by default’, emerging privacy conserving technologies and contractual controls.

Access to protected data is always strictly controlled using legally binding data sharing contracts. UKHSA welcomes data applications from organisations looking to use protected data for public health purposes. To request an application pack or discuss a request for UKHSA data you would like to submit, contact.

## Ethics statement

UKHSA have an exemption under regulation 3 of Sect. 251 of the National Health Service Act (2006) to allow identifiable patient information to be processed to diagnose, control, prevent, or recognise trends in, communicable diseases and other risks to public health.

The use of electronic health records complied with an approved Data Protection Impact Assessment, Caldicott Agreement, and Data Sharing Agreement, with approvals granted by the UKHSA Information Management and Privacy Office and the UKHSA Caldicott Office.

