## Supplementary Material for "Case Fatality Risk of Norovirus in England During a Period of Strain Replacement, 2022/23 - 2024/25 Seasons"

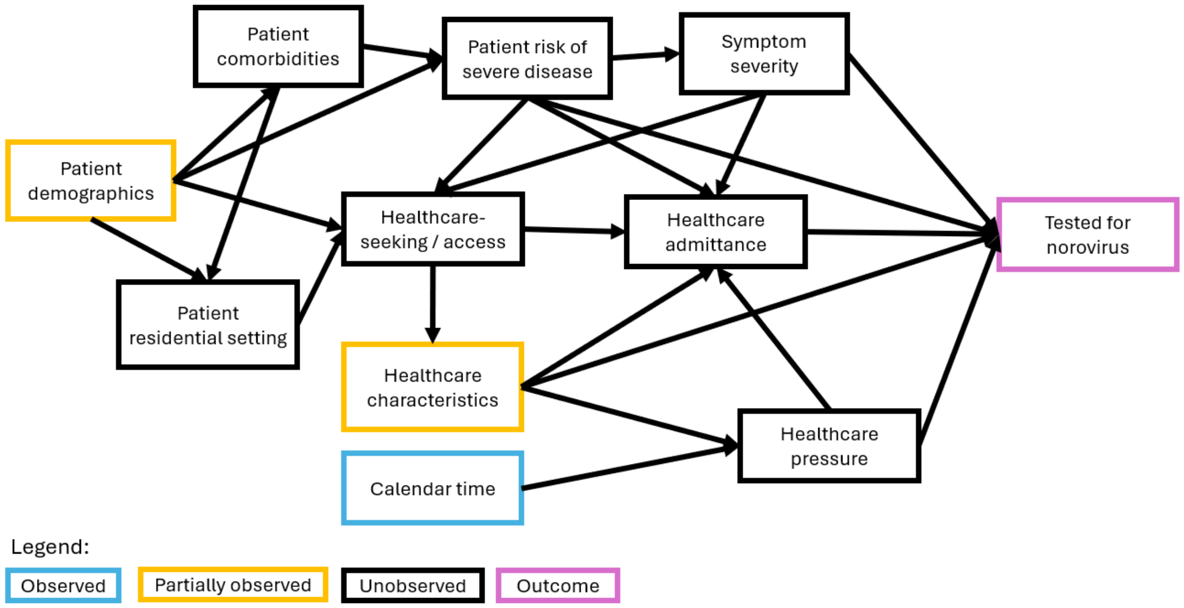


Supplementary Figure 1. Directed acyclic graph (DAG) of causal effects contributing to an individual with norovirus being tested. Observed effects are in blue, partially observed effects in orange, and the outcome in purple. Arrows show the direction of causal relationships. This does not include the decision to genotype specimens, which may have additional factors.

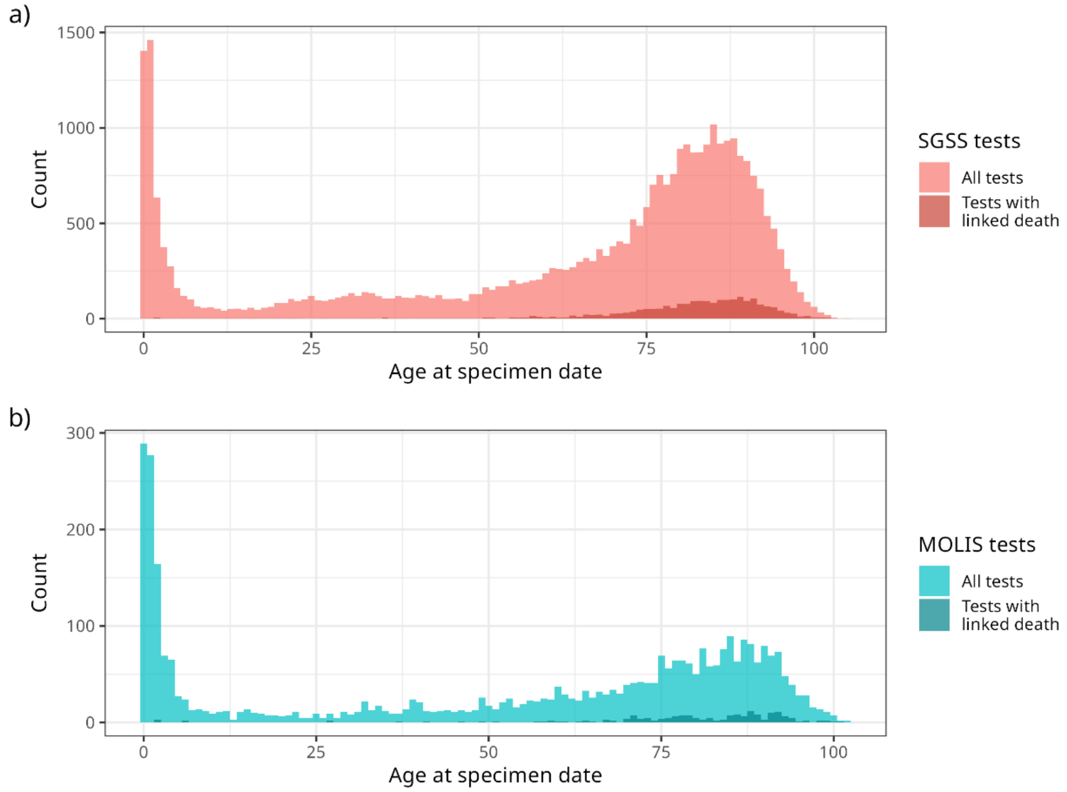


Supplementary Figure 2: Age distributions of de-duplicated test results and linked deaths within 28 days for a) **SGSS-deaths** and b) **MOLIS-deaths** data.

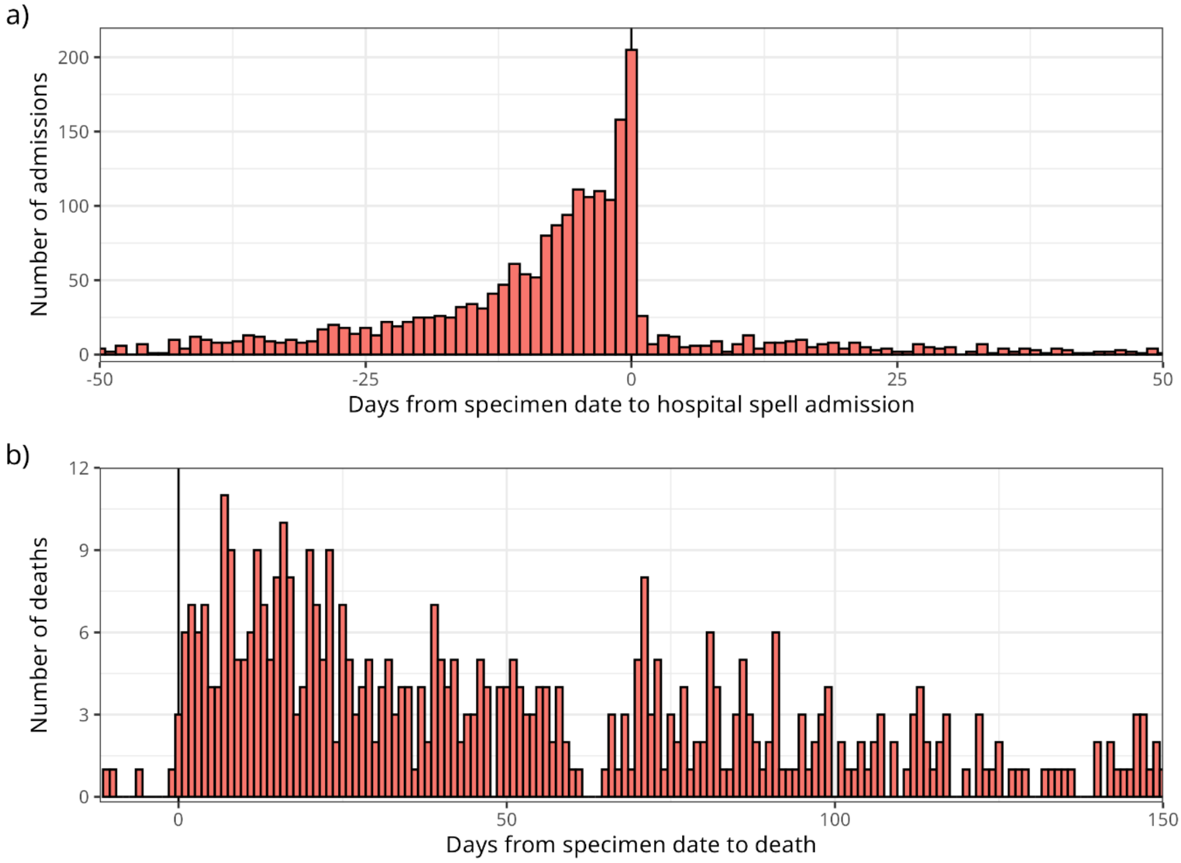


Supplementary Figure 3: For outcomes linked to a positive norovirus test from MOLIS, time from specimen date to: a) hospital spell admission date (restricted to between -50 and 50 days), and b) death (up to 150 days). A negative value of days from specimen date to outcome indicates the outcome occurring before the specimen date.

| Days from specimen to outcome date | Count | Proportion of positive tests |
|---|---|---|
| **Positive tests** | | |
| | 3542 | |
| **Hospital spell admission** | | |
| [-28, 0] | 1654 | 0.467 |
| [1,14] | 126 | 0.036 |
| [15,28] | 80 | 0.023 |
| **Hospital episode with norovirus diagnosis** | | |
| [-28, 0] | 722 | 0.204 |
| [1,14] | 35 | 0.01 |
| [15,28] | 3 | 0.001 |
| **Death** | | |
| [0,14] | 89 | 0.025 |
| [15,28] | 89 | 0.025 |
| [29,60] | 112 | 0.032 |
| [61,100] | 96 | 0.027 |

Supplementary Table 1: Summary of de-duplicated positive test results from MOLIS and linked hospital spell admissions, hospital episodes with norovirus diagnosis (ICD-10 code A081 as a primary or secondary code) or death, by linkage threshold. A negative value of days between specimen and outcome date indicates the outcome occurring before the specimen date.

| Covariate | Positive tests | Deaths | Deaths / positive tests |
|---|---|---|---|
| **Overall** | | | |
| | 1900 | 91 | 0.048 |
| **Age group** | | | |
| 0-4 | 293 | 2 | 0.007 |
| 5-19 | 89 | 1 | 0.011 |
| 20-49 | 223 | 3 | 0.013 |
| 50-74 | 391 | 13 | 0.033 |
| 75+ | 904 | 72 | 0.080 |
| **Genotype** | | | |
| GII.4 | 705 | 53 | 0.075 |
| GII.17 | 583 | 23 | 0.039 |
| Other | 519 | 9 | 0.017 |
| Unknown | 93 | 6 | 0.065 |
| **Healthcare level** | | | |
| Primary care | 266 | 4 | 0.015 |
| Secondary care | 934 | 68 | 0.073 |
| Other | 566 | 14 | 0.025 |
| Unknown | 134 | 5 | 0.037 |
| **Lab broad region** | | | |
| North | 915 | 42 | 0.046 |
| Midlands | 141 | 8 | 0.057 |
| South | 844 | 41 | 0.049 |

Supplementary Table 2: Unadjusted raw counts of de-duplicated positive test results from **MOLIS-SGSS-deaths** within 28 days by age group, genotype and healthcare level.

| Covariate | Positive tests | Deaths | Deaths / positive tests |
|---|---|---|---|
| **Overall** | | | |
| | 32427 | 2157 | 0.067 |
| **Age group** | | | |
| 0-4 | 4149 | 14 | 0.003 |
| 5-19 | 1036 | 7 | 0.007 |
| 20-49 | 3311 | 32 | 0.010 |
| 50-74 | 6856 | 354 | 0.052 |
| 75+ | 17073 | 1750 | 0.103 |
| **Healthcare level** | | | |
| Primary care | 3329 | 29 | 0.009 |
| Secondary care | 23187 | 1818 | 0.078 |
| Other | 2182 | 77 | 0.035 |
| Unknown | 3729 | 233 | 0.062 |
| **Lab region** | | | |
| East Midlands | 1272 | 101 | 0.079 |
| East of England | 2488 | 217 | 0.087 |
| London | 3059 | 98 | 0.032 |
| North East | 2927 | 190 | 0.065 |
| North West | 4775 | 324 | 0.068 |
| South East | 4015 | 255 | 0.064 |
| South West | 5387 | 403 | 0.075 |
| West Midlands | 4575 | 345 | 0.075 |
| Yorkshire and The Humber | 3929 | 224 | 0.057 |

Supplementary Table 3: Unadjusted raw counts of de-duplicated positive test results from **SGSS-deaths** data within 28 days by age group, genotype and healthcare level.

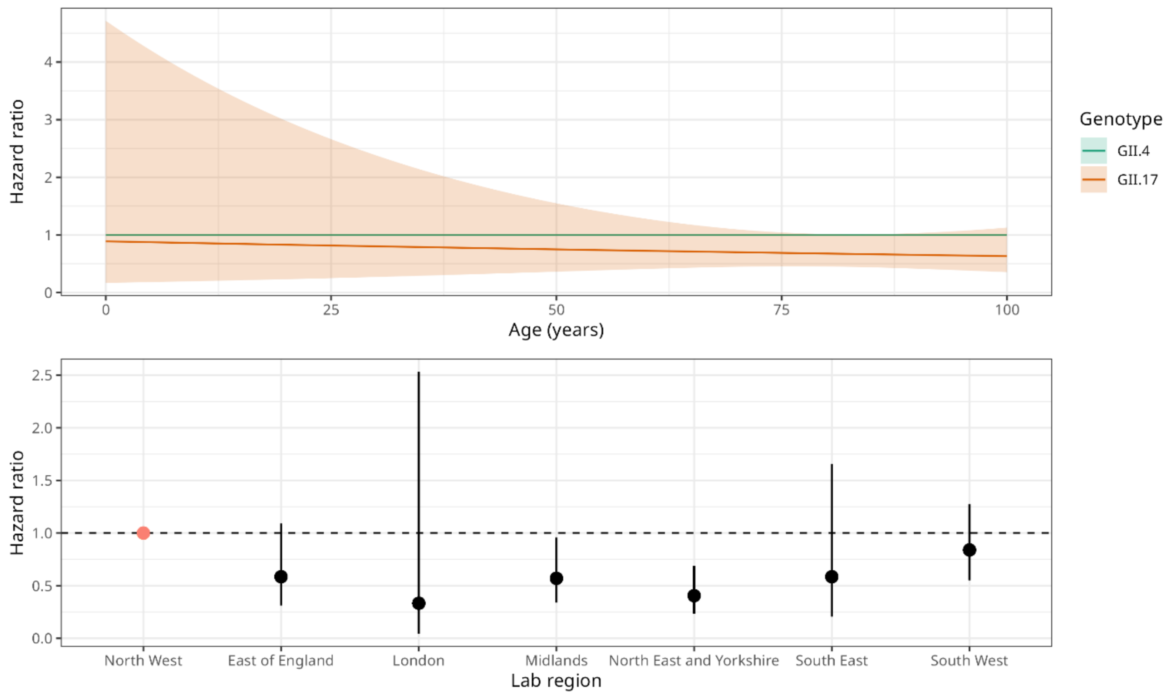


Supplementary Figure 4: Hazard ratios and 95% CIs for genotype by age and by region from CFR model using **MOLIS-deaths** data. Linkage threshold is death within 28 days. The most abundant categories were chosen as the reference genotype (GII.4) and region (North West).

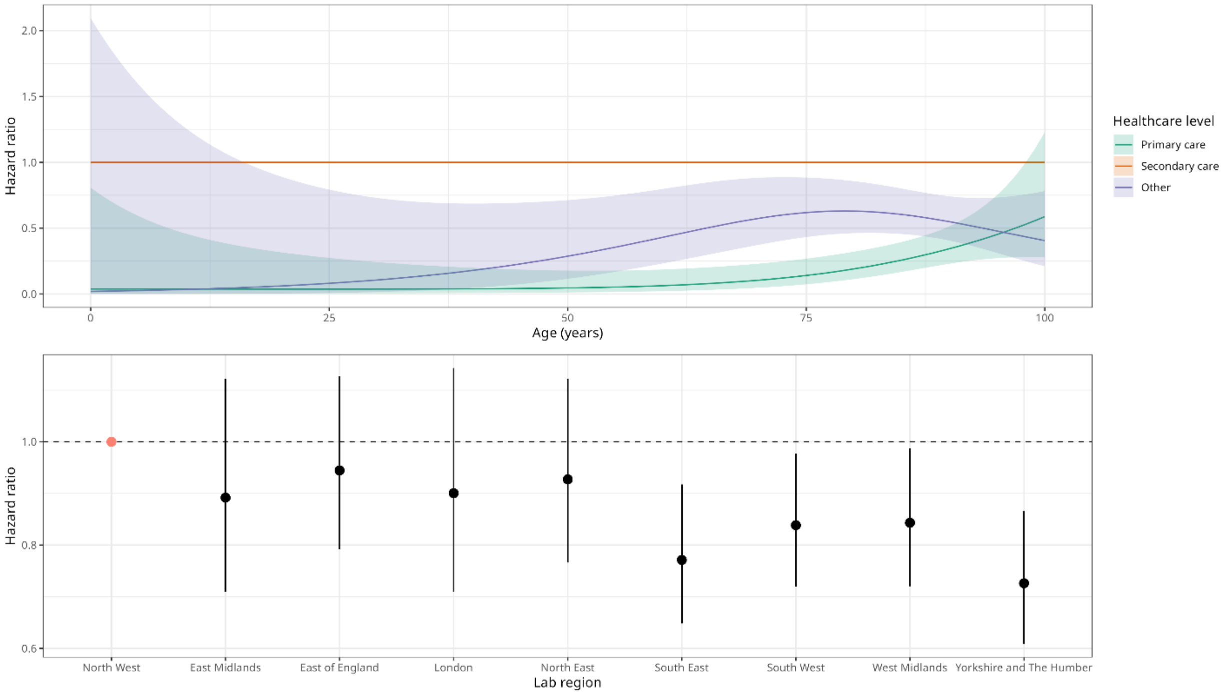


Supplementary Figure 5: Hazard ratios and 95% CIs for healthcare level by age and for lab region from CFR model using **SGSS-deaths** data. Linkage threshold is death within 28 days. The most abundant category was chosen as the reference healthcare level (Secondary care). The reference lab region (North West) was chosen to match the reference lab region in the MOLIS model.

### Supplementary Section 1: De-duplication of test result data

De-duplication of MOLIS test results retained each person's first positive test result and any subsequent positives occurring at least 21 days after the previous retained positive; however, positive results within 21 days with a different genotype from the previous result were also retained. The 21-day cutoff was chosen to align with other ongoing studies into infectious intestinal diseases and based on subject matter advice [1]. Specimens that could not be genotyped due to insufficient viral load or technical limitations were classified as "Unknown" genotype and only retained if they represented a person's sole positive test.

As with the MOLIS data, de-duplication of SGSS data retained the first positive test result and any subsequent positives occurring at least 21 days after the previous retained positive.

### Supplementary Section 2: Hospitalisation data

Hospitalisation data came from the Secondary Uses Service Admitted Patient Care (SUS APC) dataset, with both hospital spell and episode records for admissions to NHS hospitals [2]. Hospital spells represent a continuous hospital stay from admission to discharge and may consist of multiple episodes of care, each under a different consultant. Episodes carry diagnostic codes from the International statistical classification of diseases, 10th revision (ICD-10) [3]. We extracted hospital spells and norovirus-specific episodes (code A081) up to admission dates of 4 August 2025.

Hospital admissions (at the spell or episode level) were linked to the nearest positive test result in time, as tests may be taken before or after hospitalisation.

### Supplementary Section 3: Causal effects for testing

Supplementary Figure 1 shows the causal relationships contributing to someone with norovirus getting tested. Patient demographics, comorbidities and residential setting (e.g. care home) influence underlying risk of severe disease, but also healthcare-seeking behaviour and healthcare access, which contribute to probability of testing. Healthcare characteristics and system pressure further affect these and influence other factors like testing capacity and de-prioritisation of routine norovirus testing during outbreaks of other pathogens [4].

### Supplementary Section 4: Groupings of model covariates

Genotype was grouped as “GII.4”, “GII.17”, “Other” and “Unknown”. Healthcare level was grouped as “Primary care” (including only GP), “Secondary care”, “Other” and “Unknown”. Unknown values of categorical values (genotype and healthcare level) were retained so patients with missing variables could still contribute to global effects of their non-missing variables. In the MOLIS-deaths data, 368/3542 records had “unknown” genotype (Table 1). In the SGSS-deaths data, 3,729/32,427 records had “unknown” healthcare levels (Supplementary Table 3). MOLIS-deaths models used MOLIS lab regions (7 NHS regions), whereas SGSS-deaths models used SGSS lab regions (9 ONS regions). MOLIS-SGSS-deaths models used broader grouped regions (“North”, “Midlands”, “South”) due to smaller sample sizes. Records with missing age values were removed from the model data (1 or 2 records, Figure 1).

### Supplementary Section 5: MOLIS-SGSS-deaths

We can also link the MOLIS-deaths dataset to SGSS test results to get the subset of MOLIS-SGSS-deaths, which includes additional information on the healthcare level that requested the test. In the smaller MOLIS-SGSS-deaths dataset, genotypes GII.4 and GII.17 are associated with statistically similar risk of death (Supplementary Figure 6), and there is not enough statistical power to detect a difference in effect of cases tested in secondary care from primary care. The age risk profile is shown in Supplementary Figure 7, with risk roughly increasing with age, but large confidence intervals.

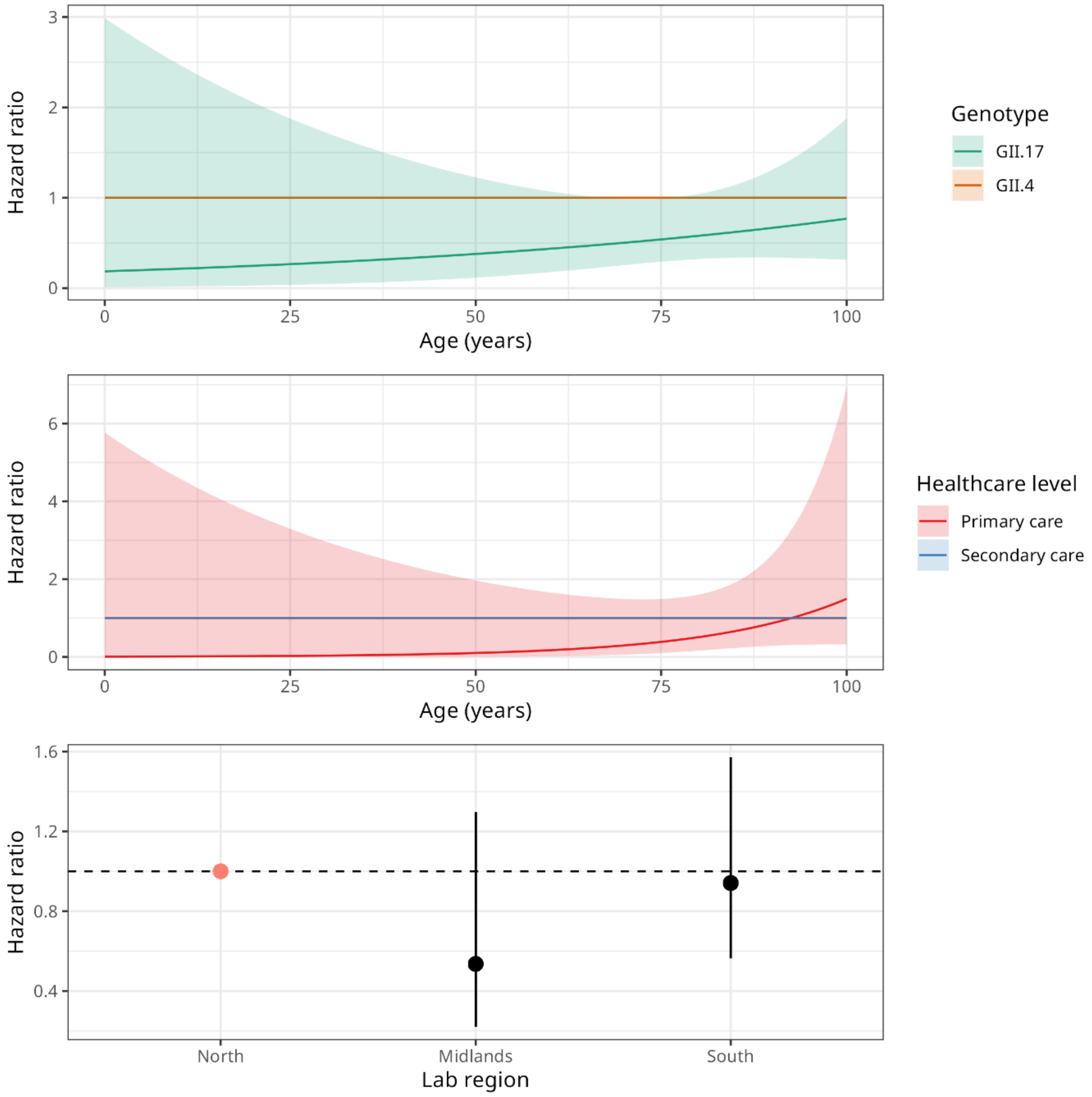


Supplementary Figure 6: Hazard ratios and 95% CIs for genotype from CFR model using **MOLIS-SGSS-deaths**. Linkage threshold is death within 28 days. The most abundant categories were chosen as the reference genotype (GII.4), healthcare level (Secondary care) and lab broad region (North).

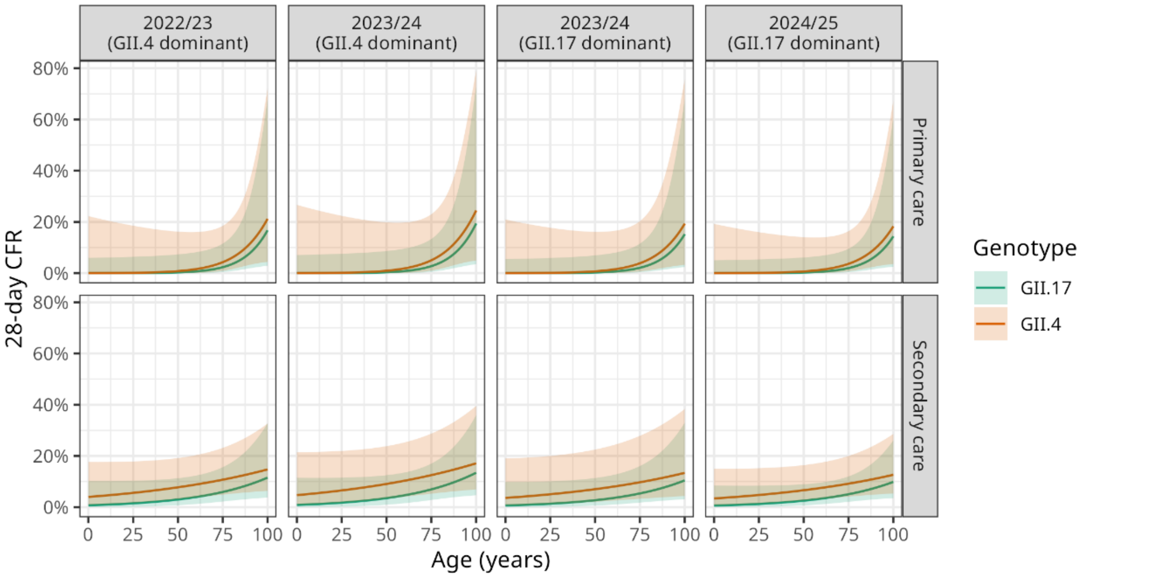


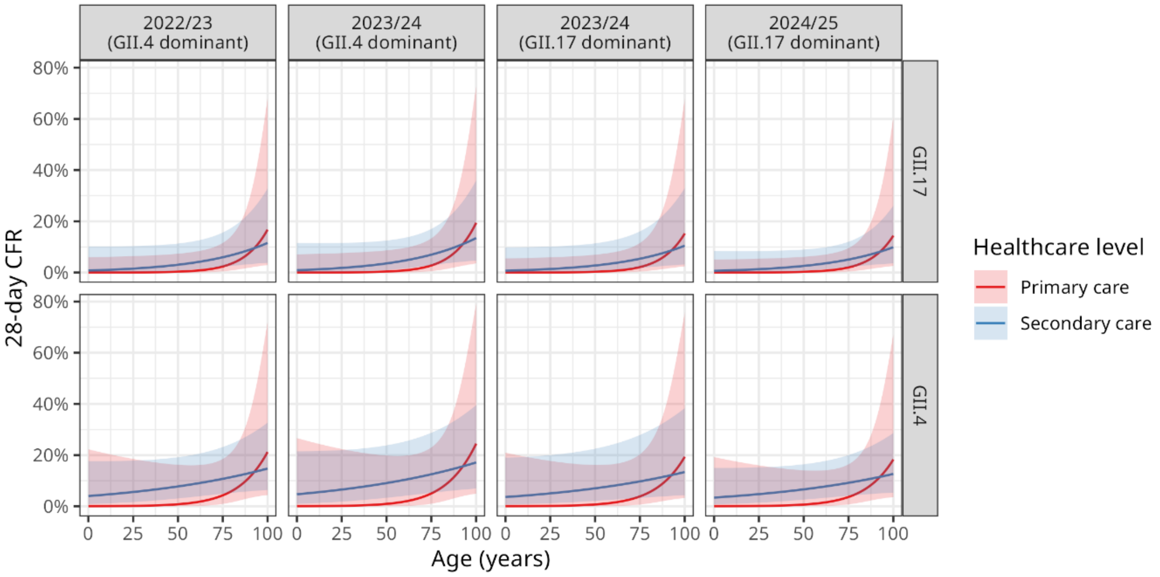


Supplementary Figure 7: Predicted age-varying case fatality risk (CFR) within 28 days of a positive test with 95% confidence intervals by genotype, time period and healthcare level using **MOLIS-SGSS-deaths** data. Time period facets were chosen to reflect periods of peak activity in each season. Two time periods are shown for the 2023/24 season to reflect periods of GII.4 or GII.17 dominance. In order, the time periods refer to test results with specimen date in the calendar quarter beginning January 2023, January 2024, April 2024 and January 2025.

### Supplementary Section 6: Sensitivity to death threshold

In the main results, the threshold for linking deaths to test results is deaths occurring within 28 days. In this section, we re-fit models changing the deaths linkage threshold to within 14 or 60 days.

#### MOLIS-deaths data

Hazard ratios for genotype and lab region for deaths within 14 and 60 days in the MOLIS-deaths data are shown in Supplementary Figure 8 and Supplementary Figure 9. GII.4 and GII.17 have statistically similar risk of death – this matches the results for deaths within 28 days.

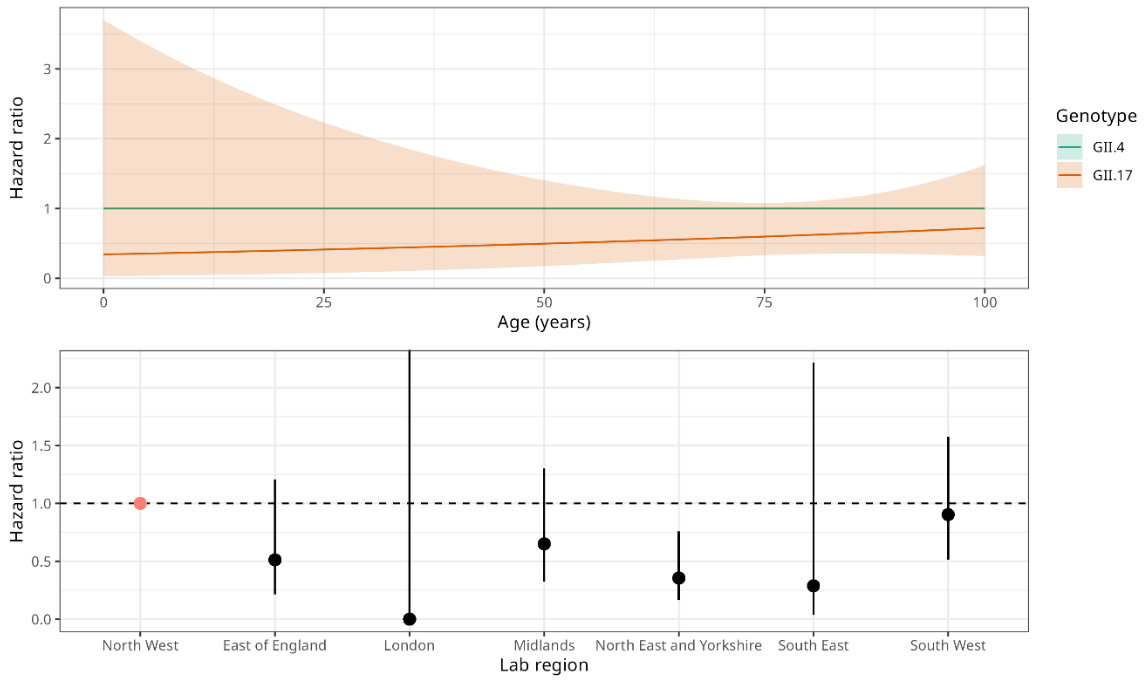


Supplementary Figure 8: Hazard ratios and 95% CIs for genotype and lab region from CFR model using **MOLIS-deaths** data. Linkage threshold is death within 14 days. The most abundant categories were chosen as the reference genotype (GII.4) and lab broad region (North West).

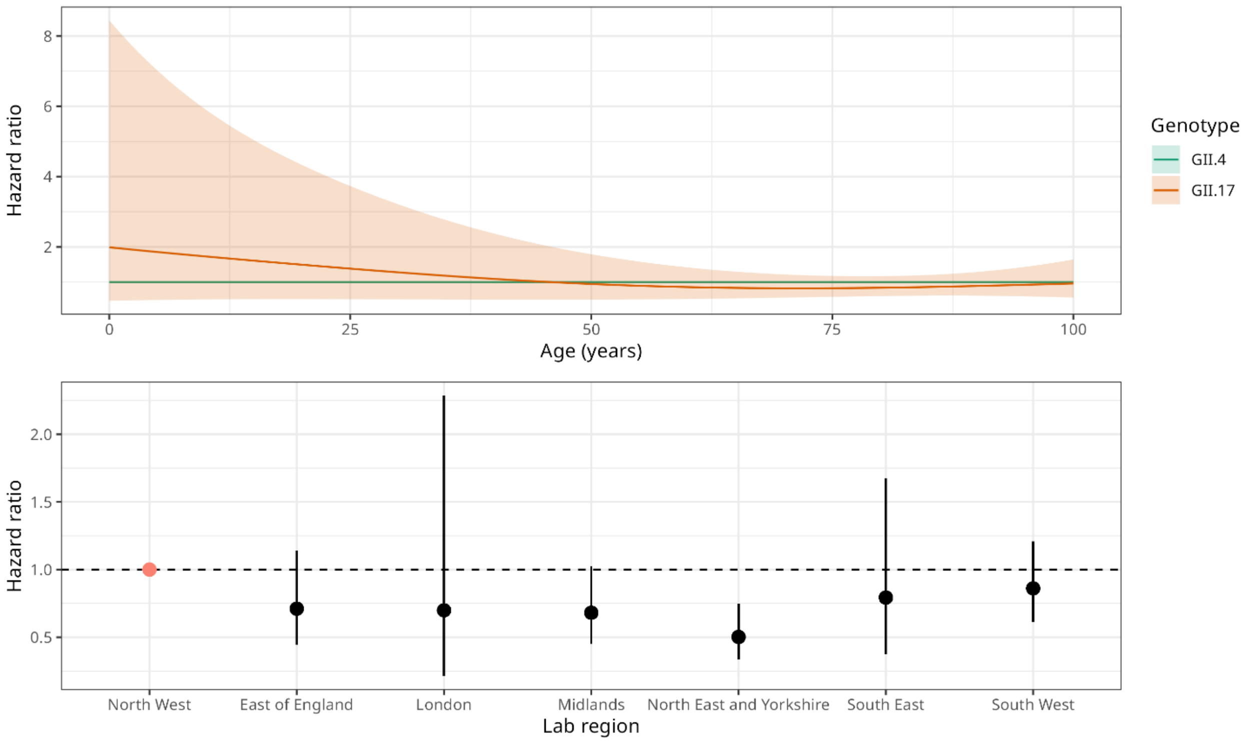


Supplementary Figure 9: Hazard ratios and 95% CIs for genotype and lab region from CFR model using **MOLIS-deaths** data. Linkage threshold is death within 60 days. The most abundant categories were chosen as the reference genotype (GII.4) and lab broad region (North West).

### SGSS-deaths data

Hazard ratios for healthcare level and lab region for deaths within 14 and 60 days in the SGSS-deaths data are shown in Supplementary Figure 10 and Supplementary Figure 11. These give similar results as deaths within 28 days. For a 14-day threshold, compared to cases identified in secondary care, those identified in primary care have significantly lower risk of death for most ages, but those in other settings have only significantly lower risk in the elderly (Supplementary Figure 10). For a 60-day threshold, compared to cases identified in secondary care, risk of death is significantly lower for cases of all ages identified in primary care or other settings (Supplementary Figure 11).

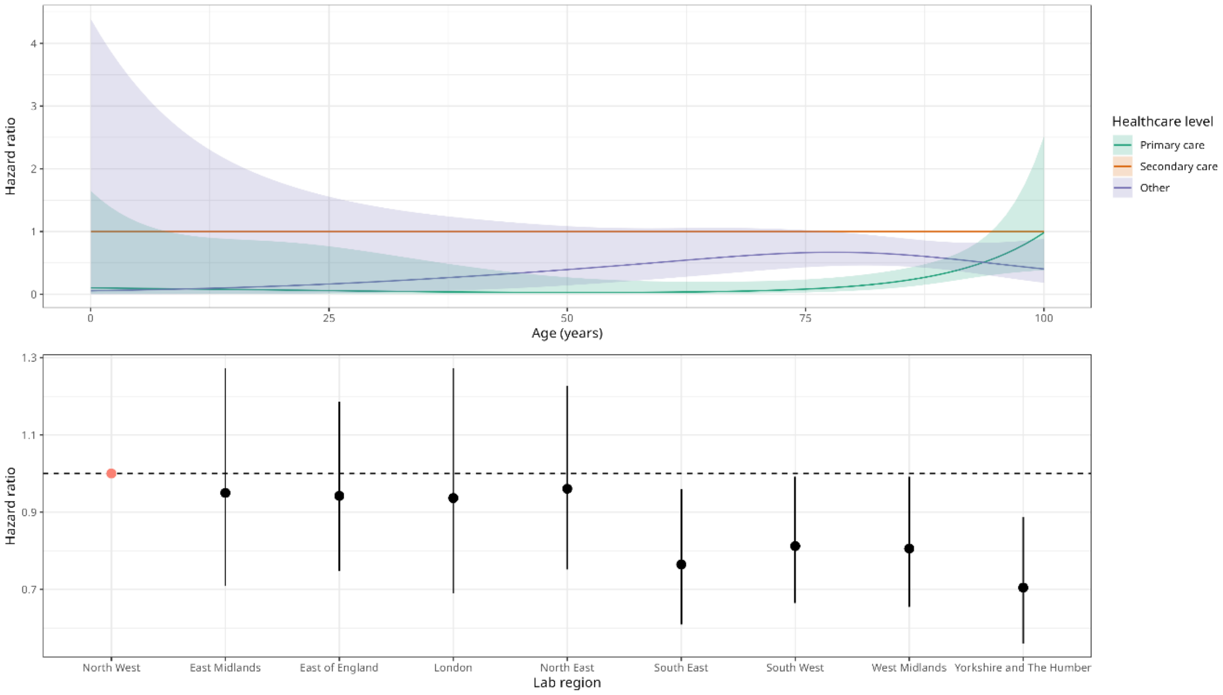


Supplementary Figure 10: Hazard ratios and 95% CIs for genotype from CFR model using **SGSS-deaths** data. Linkage threshold is death within 14 days. The most abundant category was chosen as the reference healthcare level (Secondary care). The reference lab region (North West) was chosen to match the reference lab region in the MOLIS model.

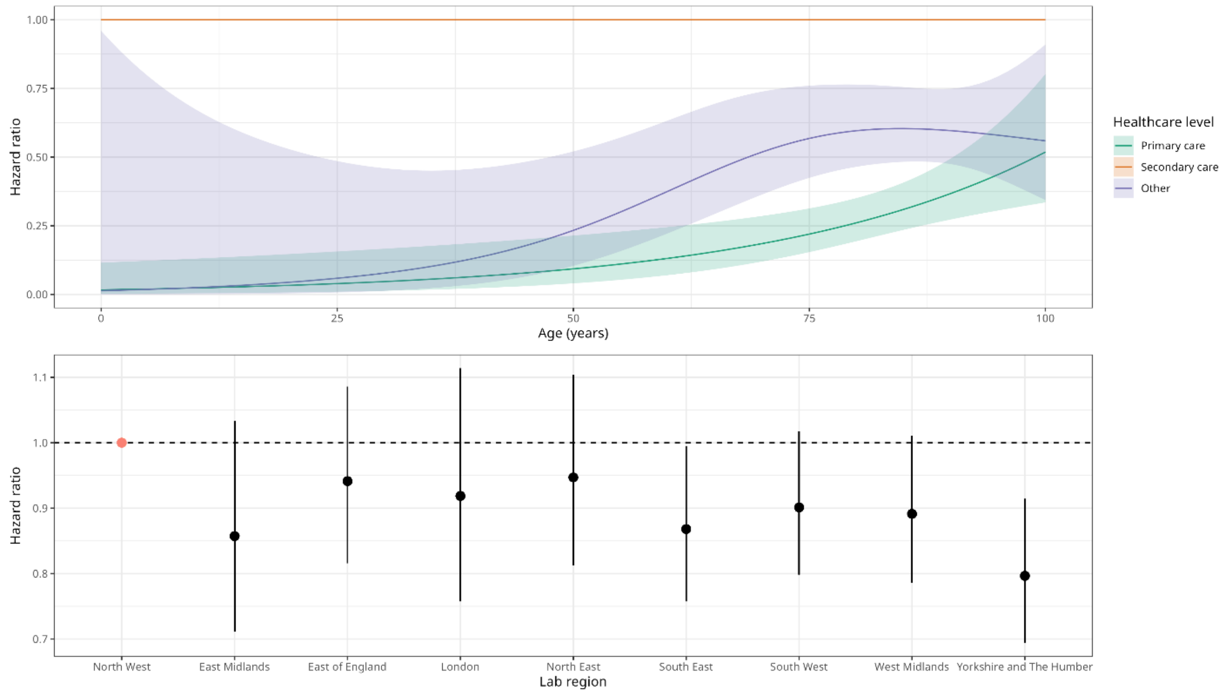


Supplementary Figure 11: Hazard ratios and 95% CIs for genotype from CFR model using **SGSS-deaths** data. Linkage threshold is death within 60 days. The most abundant category was chosen as the reference healthcare level (Secondary care). The reference lab region (North West) was chosen to match the reference lab region in the MOLIS model.

### MOLIS-SGSS-deaths data

Hazard ratios for genotype, healthcare level and lab region for deaths within 60 days in the MOLIS-SGSS-deaths data are shown in Supplementary Figure 14. We did not consider deaths within 14 days due to the small number of deaths. For deaths within 60 days, there is no statistical difference in risk between genotypes GII.4 and GII.17 and between cases identified in primary or secondary care.

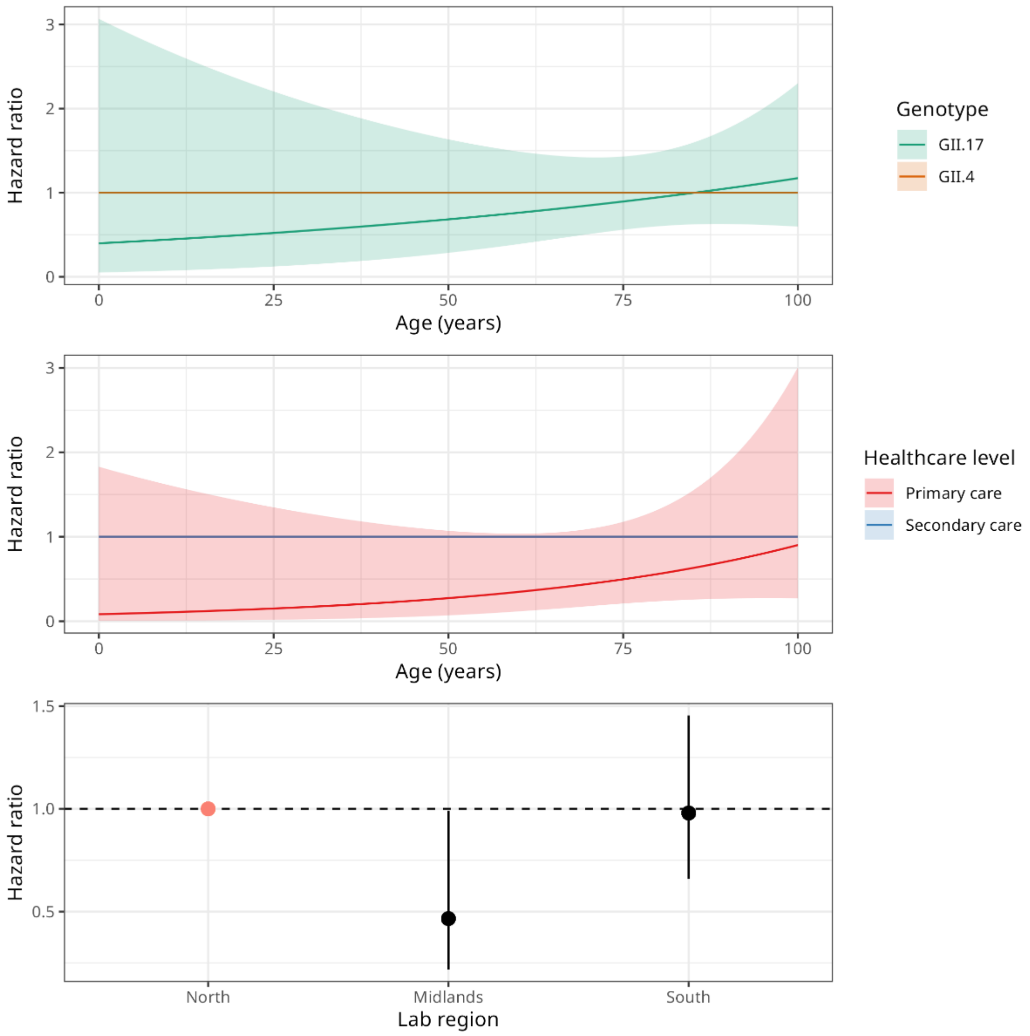


Supplementary Figure 11: Hazard ratios and 95% CIs for genotype, healthcare level from CFR model using **MOLIS-SGSS-deaths** data. Linkage threshold is death within 60 days. The most abundant categories were chosen as the reference genotype (GII.4), healthcare level (Secondary care) and lab broad region (North).
